# Sexual History Was One Of The Least Frequently Documented Social History In Cerner PowerChart Histories: A Multi-Hospital Medical Center Experience For The Year 2022

**DOI:** 10.1101/2023.11.26.23299029

**Authors:** Deepak Gupta

## Abstract

**Background:** As sexual history encompasses sensitive information about private lives, patients as well as providers may find it difficult to discuss during patient-provider encounters.

**Objectives:** To ascertain the incidence of absent documentation about sexual history among patients at seven adult hospitals of a multi-hospital medical center during the year 2022.

**Materials and Methods:** The multi-hospital medical center information technology team ran a search query in electronic medical records to extract census of unique patient identifications quantifying absent documentation rates about social history categories of Tobacco, Alcohol, Substance Abuse, Home/Environment, Sexual, Nutrition/Health, Exercise, Employment/School, Other.

**Results:** Among the patients managed in the year 2022 (n=25005), Other and Sexual social history categories were least frequently documented (1-2%). Contrarily, Home/Environment and Tobacco social history categories were most frequently documented (81-82%).

**Conclusion:** Among the patients managed at the multi-hospital medical center, sexual history was one of the least frequently documented social history during the year 2022.

## Introduction

In 2020s, society is awakening to social health parameters working in tandem for keeping patients overall healthy. History taking is an essential prerequisite during social health determination with sexual history being an important component of patients’ social history [1-7]. However, sexual history taking may be easier said than done. As sexual history encompasses sensitive information about patients’ private lives, patients as well as providers may find it difficult to discuss during patient-provider encounters. Henceforth, this may create a lag in its documentation within electronic medical records (EMR) as compared to documentation of other components in patients’ social history. Therefore, it is imperative to quantify this lag in sexual history documentation.

The purpose of this retrospective study was to ascertain the incidence of absent documentation about sexual history among patients at seven adult hospitals of a multi-hospital medical center during the year 2022.

## Materials and Methods

After Institutional Review Board approval for exempt research, the multi-hospital medical center’s information technology team ran a search query in EMR to extract census of unique patient identifications (PTIDs) depending on the completed documentation rates in their EMR’s (i.e., Cerner PowerChart) “Histories” section’s “Social History” sub-section in terms of completed “Assessment” and “Details” sub-subsections for nine “Categories”: Tobacco, Alcohol, Substance Abuse, Home/Environment, Sexual, Nutrition/Health, Exercise, Employment/School, Other.

As three adult hospitals got coalesced under the single umbrella of ZIP-Code 48201-A for the purposes of EMR search query, there were effectively only five ZIP-Coded adult hospitals during the extraction of PTIDs’ census. Thereafter, at the five ZIP-Coded adult hospitals (48201-A, 48201-B, 48201-C, 48235-D, and 48382-E), this extracted census for the year 2022 determined the following:

1. Total PTIDs At Each ZIP-Coded Hospital
2. Tobacco Section LEFT BLANK In How Many Among These PTIDs
3. Alcohol Section LEFT BLANK In How Many Among These PTIDs
4. Substance Abuse Section LEFT BLANK In How Many Among These PTIDs
5. Home/ Environment Section LEFT BLANK In How Many Among These PTIDs
6. Sexual Section LEFT BLANK In How Many Among These PTIDs
7. Nutritional/ Health Section LEFT BLANK In How Many Among These PTIDs
8. Exercise Section LEFT BLANK In How Many Among These PTIDs
9. Employment/ School Section LEFT BLANK In How Many Among These PTIDs
10. Other Section LEFT BLANK In How Many Among These PTIDs

## Results

The results of EMR search query are tabulated as percentages of total PTIDs in Table 1. Across the medical center’s adult hospitals’ PTIDs in the year 2022 (n=25005), “Other” and “Sexual” social history categories were least frequently “Assessed” with or without “Details” wherein only 1-2% PTIDs had “Other” or “Sexual” social history documented. Contrarily, “Home/Environment” and “Tobacco” social history categories were most frequently “Assessed” with or without “Details” wherein only 18-19% PTIDs had no documentation about “Home/Environment” or “Tobacco” social history. This trend in percentage prevalence of non-assessed social history categories was similar across the five ZIP-Coded hospitals of the medical center as depicted in Figure 1.

**Table 1:** Percentage Prevalence Of Non-Assessed (Left-Blank) Social History Categories Among n-Number Unique Patient IDs At Five Different ZIP-Coded Hospitals During The Year 2022.

| **YEAR 2022** | **FIVE DIFFERENT ZIP-CODED HOSPITALS** |  |  |  |  | **GRAND** |
| --- | --- | --- | --- | --- | --- | --- |
| **Social History**<br>**Category** | **48201-A**<br>**(n=7706)** | **48201-B**<br>**(n=5602)** | **48201-C**<br>**(n=2461)** | **48235-D**<br>**(n=5734)** | **48382-E**<br>**(n=3502)** | **TOTAL**<br>**(n=25005)** |
| ***Tobacco*** | 19.6% | 15.8% | 25.0% | 18.4% | 16.3% | ***18.5%*** |
| ***Alcohol*** | 67.8% | 70.7% | 75.7% | 74.7% | 64.0% | ***70.3%*** |
| ***Substance Abuse*** | 46.1% | 48.5% | 51.2% | 51.8% | 45.3% | ***48.3%*** |
| ***Home/Environment*** | 18.6% | 16.5% | 25.4% | 17.6% | 17.5% | ***18.4%*** |
| ***Sexual*** | 97.2% | 98.3% | 98.3% | 98.2% | 99.5% | ***98.1%*** |
| ***Nutrition/Health*** | 94.0% | 97.4% | 96.2% | 96.8% | 99.3% | ***96.4%*** |
| ***Exercise*** | 95.1% | 97.9% | 97.0% | 97.8% | 99.5% | ***97.1%*** |
| ***Employment/School*** | 97.0% | 98.5% | 96.5% | 97.7% | 99.3% | ***97.8%*** |
| ***Other*** | 98.9% | 99.6% | 99.6% | 99.3% | 99.8% | ***99.3%*** |

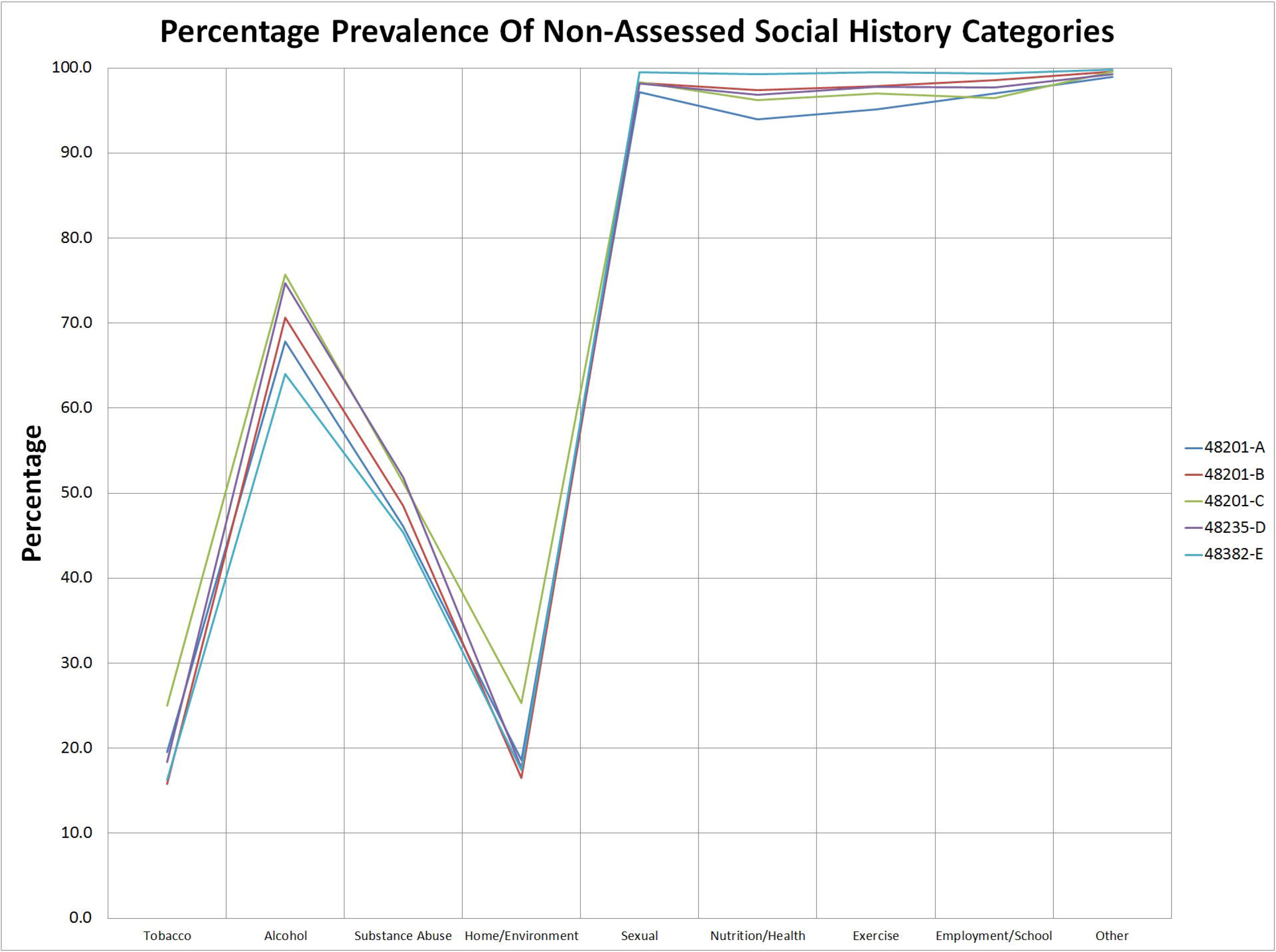

## Discussion

Gender identity spectrum and sexual orientation spectrum are vast, fluid and diversifying with implications for clinical outcomes among patients and healthcare disparities among communities. This may need objective quantification with longitudinal research into patients’ social history so as to make the case for healthcare regulators to address any issues identified during planned and envisaged current and future longitudinal research. As a good start towards this goal, the current retrospective study at a multi-hospital medical center has shown that social history in general is incompletely assessed and detailed with sexual history in particular neither routinely assessed nor routinely detailed during documentation in Cerner PowerChart Histories. The reasons for these deficiencies in EMR documentations may be many. Firstly, Cerner PowerChart Histories are traditionally completed by non-physician personnel who may not be able to comprehensively document and review all social history categories in all patients. Secondly, the clinical responsibilities may potentially make overwhelmed healthcare workers choose to document only some among the nine social history categories by potentially omitting the likes of sexual history which they may deem irrelevant for immediate patient care while covering for their potential difficulties in eliciting the likes of sexual history during high turnover patient care and management in non-private patient care areas. Finally, assuming that physicians and other writers are comprehensively documenting about all nine social history categories while writing “Physician Notes” and “PowerNotes” in Cerner PowerChart “Document Viewing”, EMR is yet to become artificially intelligent enough for auto-populating its Cerner PowerChart “Histories” section after artificially reading its “Physician Notes” and “PowerNotes”.

This study has few limitations. This retrospective study could have been extended to years beyond the year 2022 but the author could not extend this retrospective exploration due to research logistics. The documented details in social history categories could have been explored for trending the social history milieu of patients managed at the multi-hospital medical center. However, institutional review board approval had not been sought in this regard because if approved, performing the correspondingly exhaustive analysis of assessed social history details would have been logistically overwhelming for the lone researching author.

## Conclusion

Among the patients managed at the multi-hospital medical center, sexual history was one of the least frequently documented social history during the year 2022. Further research is warranted to decipher if infrequently documented sexual history is a pervasive EMR issue across the healthcare institutions globally. Henceforth, before envisaging the processes to address and overcome this deficiency, discovering the reasons for this deficiency can be a good start for healthcare researchers and healthcare administrators because none of the nine social history categories is redundantly and irrelevantly available for documentation in Cerner PowerChart Histories.

## Supporting information

EXEMPT IRB APPROVAL

## Data Availability

All data produced in the present work are contained in the manuscript

